# Physical integrity of medical exam gloves with repeated applications of disinfecting agents: evidence for extended use

**DOI:** 10.1101/2021.05.31.21258129

**Authors:** Jared S. Shless, Yoshika S. Crider, Helen O. Pitchik, Alliya S. Qazi, Ashley Styczynski, Roger LeMesurier, Daniel Haik, Laura H. Kwong, Christopher LeBoa, Arnab Bhattacharya, Youssef K. Hamidi, Robert N. Phalen

## Abstract

**Background:** The COVID-19 pandemic has created global shortages of personal protective equipment (PPE) such as medical exam gloves, forcing healthcare workers to either forgo or reuse PPE to keep themselves and patients safe from infection. In severely resource-constrained situations, limited cycles of disinfection and extended use of gloves is recommended by the U.S. Centers for Disease Control and Prevention (CDC) to conserve supplies. However, these guidelines are based on limited evidence.

**Methods:** Serial cycles of hand hygiene were performed on gloved hands using alcohol-based hand rub (ABHR) (six and ten cycles), 0.1% sodium hypochlorite (bleach) solution (ten cycles), or soap and water (ten cycles) on three types of latex and three types of nitrile medical exam gloves, purchased in the United States and India. A modified FDA-approved water-leak test was performed to evaluate glove integrity after repeated applications of these disinfecting agents. 80 gloves per disinfectant-glove type combination were tested. Within each glove type the proportion of gloves that failed the water-leak test for each disinfectant was compared to that of the control using a non-inferiority design with a non-inferiority margin of five percentage points. Results were also aggregated by glove material, and combined for overall results.

**Findings:** When aggregated by glove material, the dilute bleach exposure demonstrated the lowest difference in proportion failed between treatment and control arms: −2.5 percentage points (95% CI: −5.3 to 0.3) for nitrile, 0.6 percentage points (95% CI: −2.6 to 3.8) for non-powdered latex. For US-purchased gloves tested with six and ten applications of ABHR, the mean difference in failure risk between treatment and control gloves was within the prespecified non-inferiority margin of five percentage points or less, though some findings were inconclusive because confidence intervals extended beyond the non-inferiority margin. The aggregated difference in failure risk between treatment and control gloves was 3.5 percentage points (0.6 to 6.4) for soap and water, and 2.3 percentage points (−0.5 to 5.0) and 5.0 percentage points (1.8 to 8.2) for 10 and 6 applications of ABHR, respectively. The majority of leaks occurred in the interdigital webs (35%) and on the fingers (34%).

**Conclusion:** Current guidelines do not recommend extended use of a single-use PPE under normal supply conditions. However, our findings indicate that some combinations of glove types and disinfection methods may allow for extended use under crisis conditions. We found that ten applications of dilute bleach solution have the least impact on glove integrity, compared to repeated applications of ABHR and soap and water. However, the majority of glove and exposure combinations were inconclusive with respect to non-inferiority with a 5 percentage point non-inferiority margin. Testing specific glove and disinfectant combinations may be worthwhile for settings facing glove shortages during which extended use is necessary. The modified water-leak testing method used here is a low-resource method that could easily be reproduced in different contexts.

## Background

Medical examination gloves are an essential component of personal protective equipment (PPE) for healthcare workers (HCWs). However, in both resource-limited and crisis-capacity situations, glove supplies may be insufficient to allow for the recommended single use for every patient. The COVID-19 pandemic has strained the global supply chain for medical gloves. In March 2020, the World Health Organization (WHO) predicted that roughly 76 million medical gloves would be required each month to mount an adequate global response to the COVID-19 crisis.^1^ Manufacturers and supply chains have not kept up with demand, necessitating rationing and reuse of medical gloves as interim responses to the PPE shortage crisis.^2^ A study conducted in April-May 2020 revealed that only 36% of physicians in Pakistan had access to medical gloves.^3^ A survey of seven hospitals in Liberia during March-May 2020 found that gloves were never available in 43% of wards and were only rarely and intermittently available in 26% of wards (R. Arthur, unpublished data, 2021). These shortages endanger HCWs as well as the patients they care for.

In response to PPE shortages during the COVID-19 pandemic, the U.S. Centers for Disease Control and Prevention (CDC) published crisis capacity strategies for PPE conservation.^4^ Recommendations include extending the use of disposable medical gloves by using hand hygiene methods on gloved hands (without removing gloves) between patients or tasks, with up to six applications of alcohol-based hand rub (ABHR) or up to ten rounds of treatment with dilute bleach or soap and water.^4^ Re-use, in which gloves are taken off, disinfected, and then re-worn, is not recommended because the donning and doffing process increases the risk of tears.^4^ Since medical gloves are manufactured as single-use products, very few studies have investigated the impact of common sanitizing agents on their physical and mechanical integrity.

The CDC cites two studies to support their extended use recommendations. In the first study, researchers applied ethanol-based hand sanitizer to five brands of latex and nitrile gloves, rubbed and dried them, and filled them with an unspecified volume of water to check for leaks. Gloves were sourced from Malaysia, South Korea, and Thailand. All brands of gloves were leak-free after 30 applications of ethanol-based hand sanitizer, and one brand of latex gloves was leak-free after 100 cycles of disinfection with 83% ethanol solution.^5^ Although the study was limited by a small sample size (ten gloves of each brand), the results indicated that glove integrity may be maintained well beyond six disinfection cycles. The second study showed that latex and some nitrile gloves could be disinfected with ethanol-based ABHR up to six times without any significant change in tensile strength.^6^ For the recommendation to use dilute bleach solution as a glove disinfectant, the CDC cites a document from a glove manufacturer that reported no detectable permeation in tests of its nitrile gloves using 10-13% sodium hypochlorite.^7^ It is not known how these results from a single manufacturer apply to other glove brands or materials. No published reports were found on the effect of soap and water on glove integrity.

This study aimed to evaluate the effect of repeated applications of ABHR, dilute bleach, and soap and water on medical exam glove integrity. To ensure these results may be used in high-and low-resource settings, we sourced latex and nitrile gloves from both the U.S. and India.

## Methods

### Gloves

Three types of nitrile gloves and three types of latex gloves were selected for testing (Table 1). All gloves were single-use medical exam type gloves. Three glove types were commercially purchased in the United States: Glovepak nitrile (Glovepak USA, Palm Desert, CA, USA), Polymed latex (Ventyv, Tampa, FL, USA), and SemperSure nitrile (Sempermed, Clearwater, FL, USA). Three glove types were commercially purchased in India and shipped to the U.S. for testing: Surgi Gloves powdered latex, Surgi Gloves nitrile, and Surgi Gloves non-powdered latex (Connect Pack LLP, Mumbai, India). In 2016, the U.S. Food and Drug Administration (FDA) banned the use of powdered examination gloves due to risk of allergic reactions, yet in low- and middle-income countries such as India, these gloves are still widely used.^8,9^ According to the packaging, the Polymed latex gloves were manufactured in Thailand; the rest were manufactured in Malaysia. For four of the six glove types, glove testers selected the size that best fit their hand (medium or large). Average glove thickness at the palm ranged from 0.055 mm (Glovepak nitrile) to 0.103 mm (Polymed latex) (Table 1). Given visibly different glove quality between sizes (e.g., damaged gloves out of the box, different thicknesses), only medium sized Surgi Gloves non-powdered latex and large sized Surgi Gloves nitrile gloves were tested.

**Table 1:** Summary of gloves tested.

| Material | Brand | Sizes | Water-leak test volume [L] (size) | Country of purchase | Country of manufacture | Glove palm thickness [mm] |
| --- | --- | --- | --- | --- | --- | --- |
| Latex | Polymed® non-powdered | Med/Lg | 2 (Med)<br>2.5 (Lg) | U.S. | Thailand | 0.103± 0.011 |
|  | Surgi Gloves® powdered | Med/Lg | 1.5 | India | Malaysia | 0.076± 0.011 |
|  | Surgi Gloves® non-powdered | Med | 2.5 | India | Malaysia | 0.095± 0.014 |
| Nitrile | SemperSure® | Med/Lg | 1.5 | U.S. | Malaysia | 0.062± 0.004 |
|  | Glovepak® Europa | Med/Lg | 1 | U.S. | Malaysia | 0.055± 0.003 |
|  | Surgi Gloves® | Lg | 1.5 | India | Malaysia | 0.059± 0.005 |

### General setup and calibration: modified water-leak test

For each exposure method, we employed a modified FDA water-leak test, in which leaks are detected by filling gloves with water and observing any leaks that form over two minutes. The original method uses a standard 1L water volume.^10,11^ The modified water-leak test includes an initial step to calibrate the water volume to each glove type to increase the sensitivity of the test, as compared to the FDA standard method, to detect small defects that could pass a virus.^12,13^ The water volume used for the water-leak test for a specified glove type was based on the minimum volume necessary to detect visible leaking from 30-gauge needle punctures spaced across the glove (see additional details in Supplementary Materials).^12,14^ Each glove type and size were individually calibrated (Table 1).

To perform the water-leak test, each glove was attached to the base of a Schedule 40 PVC pipe that was two inches in diameter and two feet long (Figure 1). The edge of the wrist of the glove was positioned four centimeters from the end of the pipe and secured with a PVC coupling; the other end of the pipe was suspended by a wire from a hanging rack. The calibrated water volume was slowly added to the top of the column and a timer was set for two minutes. After two minutes, the glove was recorded as a “pass” if no leaks were observed through visual and tactile inspection, or a “fail” if any leaks were observed during the two minute test period. The locations of visible leaks were recorded.

**Figure 1:**
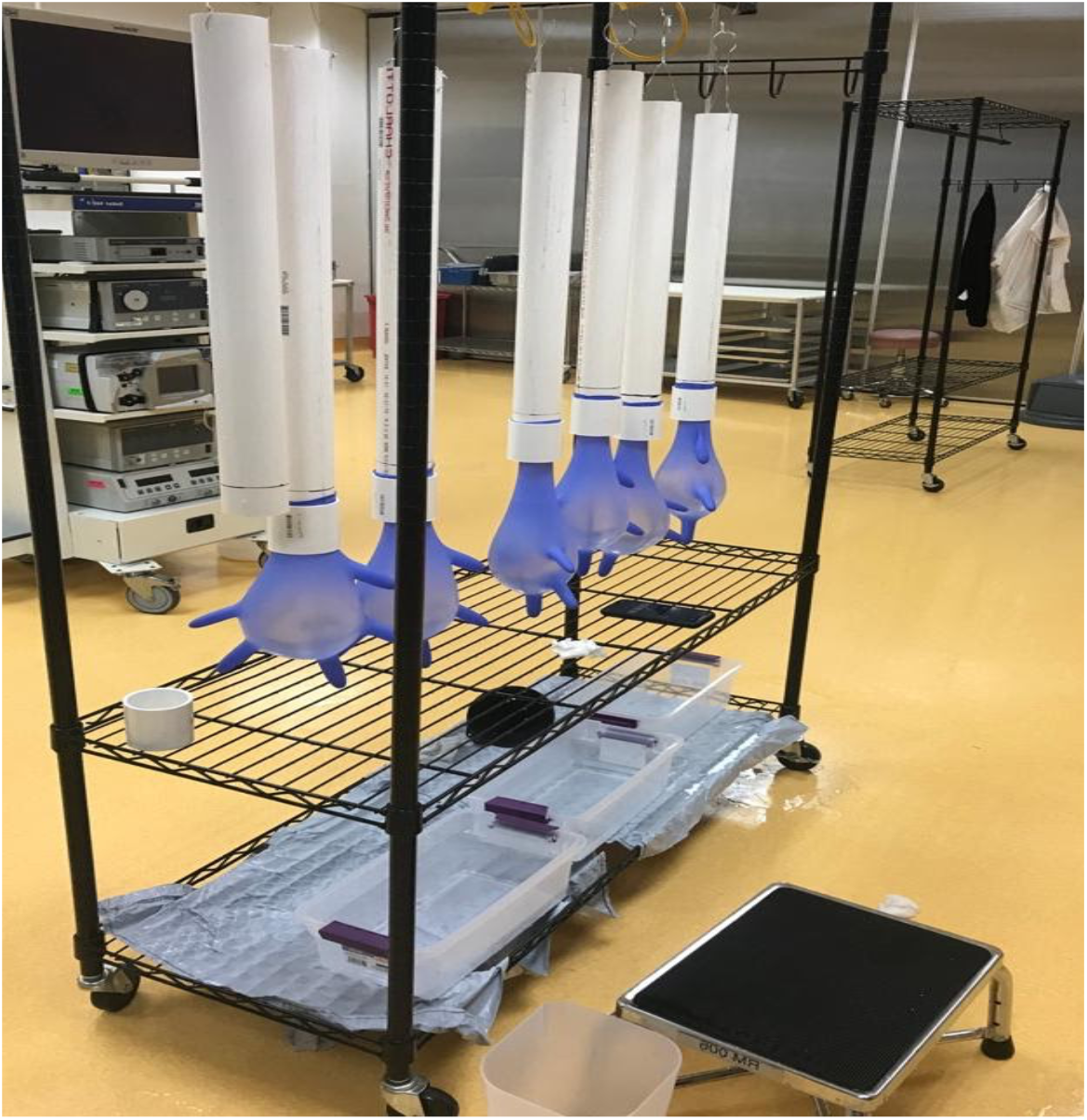
Water-leak test set-up. Figure note: Set up to test physical integrity of gloves using the water-leak method

### Exposure

Prior to the exposure, a glove was discarded if it had any visible defects or tears. The study team removed any rings from their fingers prior to donning gloves and conducting the following exposure procedures. Sanitizing procedures were adapted from recommendations by the CDC and WHO (Supplementary Table 1).

### Control

In the control arm, gloves were donned and doffed one time to simulate mechanical stress associated with typical glove use. No further treatments or manipulation were applied prior to conducting the water-leak test.

### Alcohol-Based Hand Rub (ABHR)

One full pump of Purell® Advanced Gel Hand Sanitizer (70% ethyl alcohol, GOJO Industries Inc, Akron, OH) was dispensed onto gloved hands. Gloved hands were rubbed together, palm-to-palm, working the ABHR evenly over the surface of both hands in accordance with CDC and WHO guidance for hand hygiene (Supplementary Table 1).^15,16^ The total time for each application was 20 seconds. Gloved hands were held up to dry for 20 seconds before proceeding with the next ABHR exposure or removing for water-leak testing. For ABHR, six exposures (ABHR-6) and ten exposures (ABHR-10) were separately performed.

### Dilute bleach solution

To facilitate testing, racks holding multiple pairs of gloves were constructed using wire and clothespins (Supplementary Figure 1). Gloves were donned and doffed prior to attaching it to the rack. Each rack of gloves was dipped into a prepared solution of 0.10% sodium hypochlorite (The Clorox Company, Oakland, CA) for five seconds, ensuring complete coverage on the exterior of gloves and taking care to keep wrist openings above the solution. The racks were then held above the bleach solution without rinsing for one minute to ensure adequate contact time for disinfection.^4^ Gloves were maintained in a downward position, with fingertips facing the bleach solution, to avoid getting bleach inside gloves. To rinse excess bleach, the glove racks were gently dipped into a bucket of clean, room-temperature water, again taking care to keep wrist openings above the solution. Gloves were then gently shaken to remove any excess water and dried with a paper towel before proceeding with the next dilute bleach exposure or removing for water-leak testing. Ten repeated exposures to dilute bleach solution were conducted.

### Soap and Water

First, gloved hands were wetted with room-temperature water. Next, a single pump of Dial® Basics Hypoallergenic liquid hand soap (Henkel Consumer Goods Inc, Stamford, CT) was applied to the palm, and hands were rubbed together, working the soap and water evenly over the surface of both hands for 40 seconds, in accordance with CDC and WHO techniques for hand hygiene (Supplementary Table 1).^15,16^ Gloved hands were rinsed thoroughly with room-temperature water before drying with a single-use paper towel. Ten repeated exposures to soap and water were performed.

### Statistical Analysis

The sample size was based on a non-inferiority trial design, comparing a glove exposed to a sanitizing procedure to an unexposed control glove. The primary outcome was the proportion of gloves that failed the modified FDA water-leak test. Based on a conservative assumption of a 2.5% risk of failure (the acceptable FDA rejection limit for medical gloves using the standard water-leak test) among control gloves, a five percentage point non-inferiority margin, and 90% power, at least 75 gloves per exposure-glove combination was required. A slightly higher final sample size of 80 gloves was selected to align with published sample size tables in FDA guidance for glove testing.^14^ Non-inferiority is demonstrated when the confidence interval for the difference between the treatment and the control is entirely below the non-inferiority margin. A five percentage point non-inferiority margin (the percent of failures in the treated gloves minus the percent of failures in the control gloves not exceeding five percentage points) was used, based on the 2.5% FDA threshold and a non-inferiority margin deemed reasonable according to a prior study that evaluated 30 different glove brands with the out-of-box percentage of failures ranging from 0% to 6.25%.^12^

## Results

### Risk of failure by treatment method

Risk of failure in the control gloves ranged from 1.3% to 11.3% (Table 1, Supplementary Figure 2). Two of the six glove types, both from India, exceeded the *a priori* expectation of 2.5% risk of failure in the control group. Pooling by glove material within the control arm, resulted in a slightly higher failure risk among latex gloves (5.0%, 95% confidence interval [CI]: 2.2-7.8) compared to nitrile gloves (3.8%, 95% CI: 1.3-6.2), which includes the 11.3% failure risk observed for powdered latex gloves. Among non-powdered latex gloves, the observed failure risk in the control arm was 1.9% (95% CI: 0.0-4.0). Among both latex and nitrile glove types, the bleach exposure demonstrated the lowest failure risk: 5.4% (95% CI: 2.6-8.3) for all latex and 1.3% (95% CI: 0.0-2.7) for all nitrile. The repeated soap and water exposure led to the highest risk of observed failures: 14.2% (95% CI: 9.8-18.6) for all latex and 6.7% (95% CI: 3.5-9.8) for all nitrile.

**Table 2:**
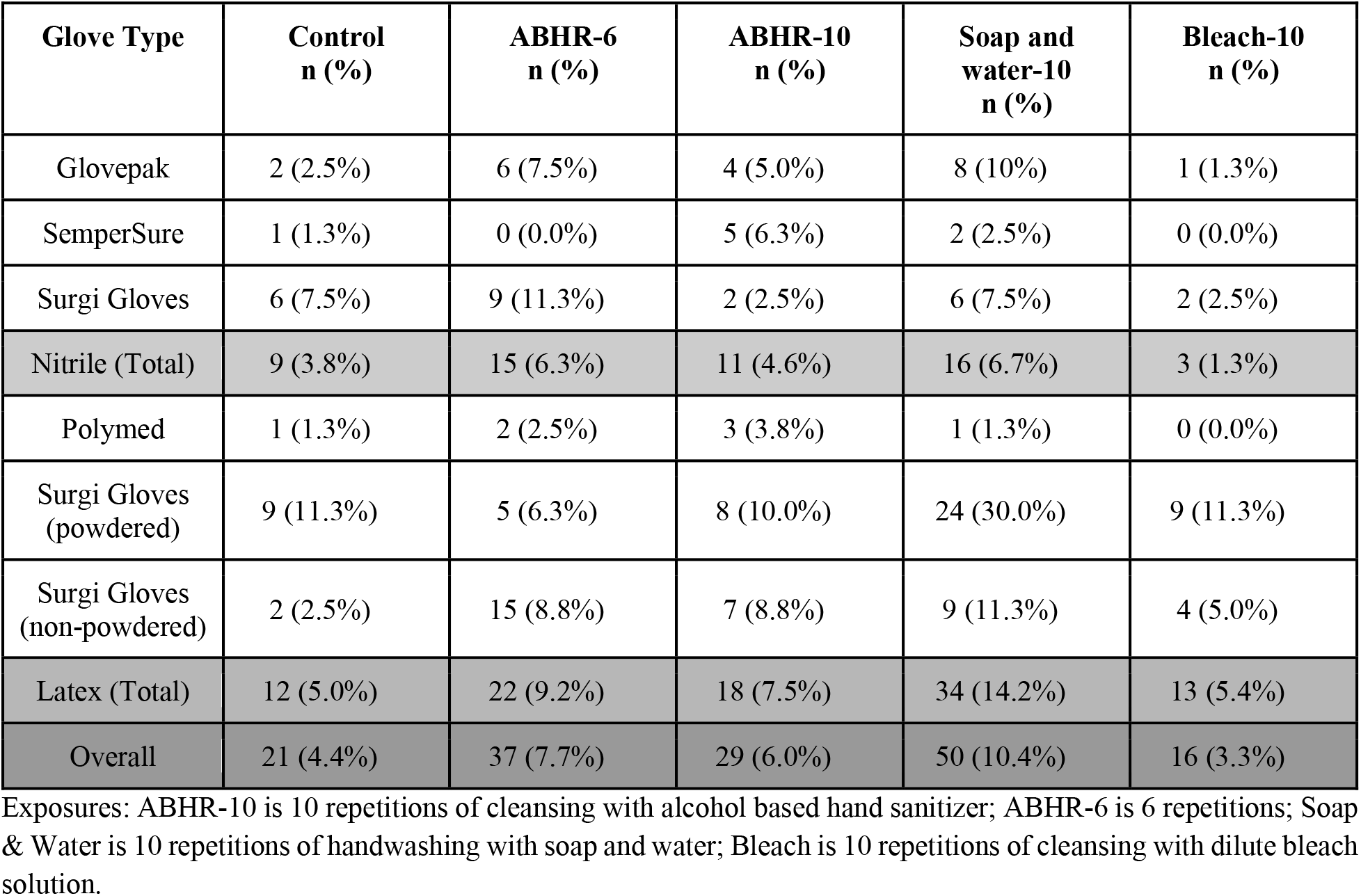
Glove leakage failures after exposure to sanitizing agents.

### Difference between treatment and control

Four of the six glove types demonstrated non-inferiority when comparing gloves treated with ten applications of bleach to their control gloves. The two glove types that did not demonstrate non-inferiority were both latex glove types from India (Figure 2). Two of the six glove types demonstrated non-inferiority when treated with six applications of ABHR (Figure 2). A single glove type showed non-inferiority with ten applications of ABHR but did not meet this criterion for six applications. Only one glove type met non-inferiority for ten applications of soap and water (Figure 2).

**Figure 2:**
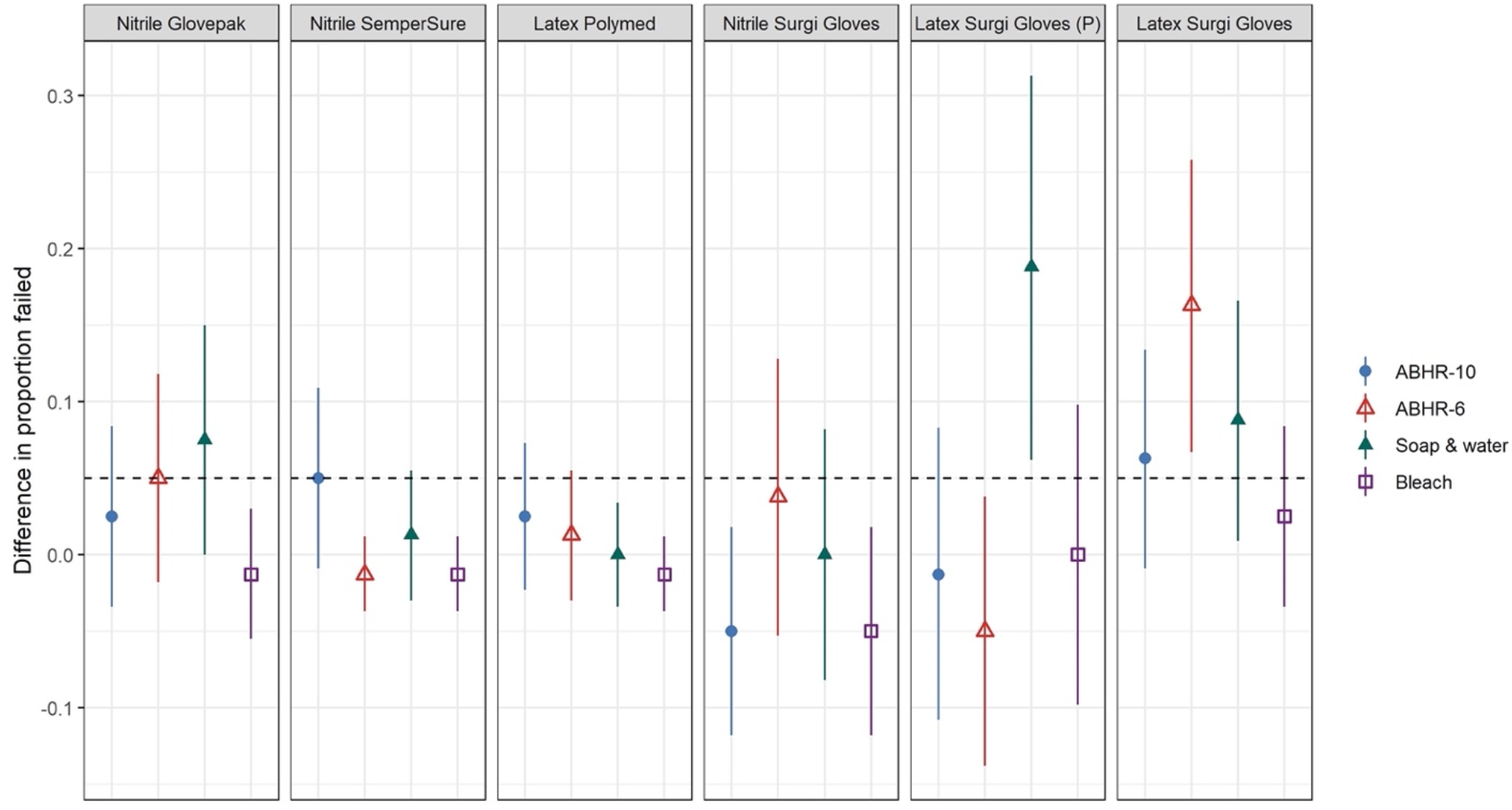
Differences in proportions of gloves that failed the water-leak test between treatment and control arms. Figure notes: The left three panels show gloves purchased in the U.S. The right three panels show gloves purchased in India. Water-leak test results are shown as points that represent differences in the proportion of glove failures between treatment and control (treatment minus control). Bars represent the 95% confidence interval for the difference. The non-inferiority margin was 0.05, represented by the dashed line. Non-inferiority is established if the confidence interval is fully contained below the non-inferiority margin. Sample size was n=80 for each arm. P=powdered (all other gloves non-powdered) Exposures: ABHR-10 is 10 repetitions of cleansing with alcohol based hand sanitizer; ABHR-6 is 6 repetitions; Soap & Water is 10 repetitions of handwashing with soap and water; Bleach is 10 repetitions of cleansing with dilute bleach solution

The majority of glove and exposure combinations were inconclusive (5/6 glove types for ABHR-10, 4/6 for ABHR-6, 4/6 for soap and water, 2/6 for bleach), meaning that the confidence interval crossed the non-inferiority margin (Figure 2). Two combinations of exposures and glove types from India demonstrated inferiority (the entire confidence interval for the difference lies outside of the non-inferiority margin): 1) the powdered latex glove exposed to soap and water and 2) the non-powdered latex glove exposed to six applications of ABHR (Figure 2). However, the same non-powdered latex glove type exposed to ten applications of ABHR has an inconclusive result.

When aggregating data by glove material, and excluding the powdered latex gloves, the mean difference in the risk of glove failure between control and exposed gloves was largest for six applications of ABHR on latex gloves (8.8 percentage points, 95% CI: 3.4-14.1) (Figure 3). Failure risk was lowest for bleach on nitrile gloves (−2.5 percentage points, 95% CI: −5.3 to 0.03) (Figure 3).

**Figure 3:**
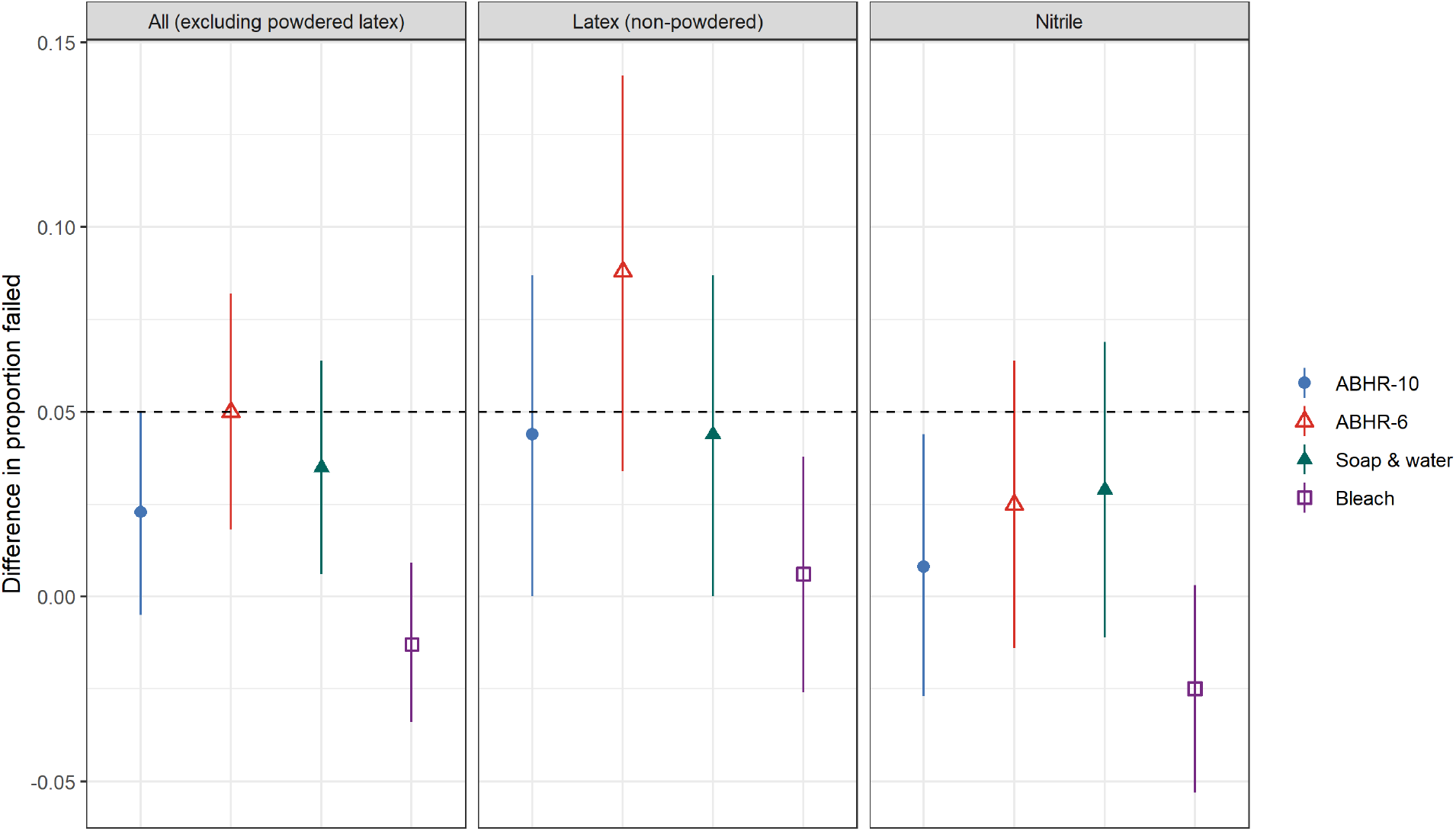
Aggregated differences in proportion failed by exposure and glove material (excluding powdered gloves) Figure notes: Water-leak test results are shown as differences in the proportion of glove failures between treatment and control (treatment minus control), excluding the latex powdered gloves. The bar represents the 95% confidence interval for the difference. The dashed line represents the non-inferiority margin (0.05). The total sample size was n=400, with n=160 for latex gloves (excluding 80 latex powdered gloves) and n=240 for nitrile gloves. Exposures: ABHR-10 is 10 repetitions of cleansing with alcohol based hand sanitizer; ABHR-6 is 6 repetitions; Soap & Water is 10 repetitions of handwashing with soap and water; Bleach is 10 repetitions of cleansing with dilute bleach solution

### Water-leak failures by glove location

There were 153 gloves that failed the water-leak test out of 2400 gloves tested. Glove failures were categorized based on leak location on the glove, including both the exposed and control gloves tested (Supplementary Table 2). 207 individual leaks were recorded. The majority of leaks occurred in the interdigital webs (35%) and on the fingers (34%), with 4% of these failures occurring at the fingertips and 30% observed elsewhere on the fingers. The palms of the gloves sustained 24% of total failures. A small number of failures could not be categorized, with 4% of gloves bursting entirely and 3% undetermined due to recording error.

A large number of Surgi Gloves nitrile gloves were discarded before testing due to visible defects such as thinning, discoloration, and tears. CDC guidelines for hand hygiene on gloved hands recommend discarding any gloves with visible defects.^4^

## Discussion

Evidence-based recommendations are provided to inform the disinfection and extended use of latex and nitrile medical gloves in crisis capacity scenarios. Notably, we found wide variability in quality and performance across gloves purchased in the U.S. and India. Even though powdered latex gloves were included in the experiment due to their continued use in many settings, they were ultimately excluded from the aggregate analysis. This decision was based on recommendations against their use in the medical setting, in addition to our finding that they demonstrated the highest risk of failure among all the glove types, with a control arm failure risk of 11.3%, or 1.5-8.7 times the observed failure risk of other tested gloves.^8,17^

Across all latex and nitrile gloves, sanitizing with dilute bleach resulted in the lowest failure risk. The results show that nitrile gloves may be cleaned with dilute bleach solution ten times without increasing the risk of tears or holes by more than five percentage points (i.e., demonstrating non-inferiority). Latex gloves purchased in the US also maintained their integrity after sanitizing with bleach, demonstrating non-inferiority. The results for both types of latex gloves purchased in India were inconclusive, although the mean difference in failure risk was less than five percentage points for the non-powdered latex gloves.

For six and ten applications of ABHR, the mean difference in failure risk was five percentage points or less for the latex and nitrile gloves purchased in the US, although some confidence intervals extended above the non-inferiority margin. This suggests that up to ten applications of ABHR may only moderately increase the likelihood of tears or holes. In aggregate, ten applications but not six applications of ABHR met non-inferiority criteria. 29 gloves failed the water-leak test after exposure to ten applications of ABHR versus 37 gloves that failed after exposure to six applications of ABHR, demonstrating that conclusions of non-inferiority in the study are sensitive to relatively small changes in the number of failures. The results presented here differ from a previous study finding that all of the latex and nitrile glove brands tested could be sanitized with ABHR up to 30 times without any gloves failing a water-leak test.^5^ However the previous study used an unspecified volume of water for water-leak tests and only ten gloves per type, which provided an unknown sensitivity to detect small holes and very little power to detect leaks. The thorough hand hygiene procedures used in this study may also have caused more wear on gloves than in past studies.

Sanitizing gloves with soap and water demonstrated the widest range of failure risks when comparing glove types, making it the least consistent disinfection option. Notably, the choice of a five percentage point non-inferiority margin is conservative as prior studies have found up to 6.25% failure risk among out-of-the-box gloves.^12^ Additionally, a higher failure risk was expected with testing gloves using a more sensitive modified water-leak test as compared to the standard 1 L FDA water-leak test.

There are some practical considerations in selecting a disinfection protocol for extended use of gloves. For example, HCWs may consider it impractical to decontaminate gloves with bleach because of the one-minute wait required for sufficient disinfection contact time. It was also observed that during 40-second soap and water treatments, water may seep into the gloves at the wrist. CDC thus recommends wearing longer cuffed gloves for disinfection by either soap and water or dilute bleach solution.^4^ Where those gloves are unavailable, an alternative may be to seal gloves at the wrist with medical tape. Additionally, gloves treated with ABHR were observed to become tacky or sticky. These observations were similar to those made by Gao et al. (2016), who recommended that HCWs test their disinfectant options with the type of gloves they will use, in case changes such as stickiness render certain tasks more difficult. In terms of convenience, ABHR is the fastest disinfection option for gloved hands.^6^

The strengths of this study are that it employs a simple water-leak testing method that uses widely available, low-cost materials, and it can be easily reproduced in a variety of settings. In addition, we included common disinfectants manufactured by global companies that distribute the same or similar products internationally. Finally, we included gloves sourced from both India and the U.S., which allowed an evaluation of the effect of disinfection on glove integrity for gloves that are used worldwide and may not meet U.S. regulatory standards.

The study has some limitations. The gloves purchased in India were sourced from a single company, so they may not be representative of the variety available for purchase on the market. Additionally, since the recommended CDC and WHO protocols for disinfection of gloves/hand hygiene were strictly followed, the resulting exposures may have been longer or more physically abrasive to the gloves than real-life hand hygiene behaviors, which may be performed for less time or with less intensity than recommended. Finally, although the non-inferiority margin was selected based on the FDA guidance, evidence in the literature, and expert opinion, there is no standard accepted margin for increased risk of tears and leaks in medical examination gloves.

In conclusion, wide variability was observed in the effect of disinfecting latex and nitrile medical examination gloves with repeated applications of ABHR, dilute bleach solution, and soap and water. The findings support the recommendation of glove disinfection with ten applications of dilute bleach solution in settings with severe PPE shortages. ABHR and soap and water had mixed effects depending on the glove type. Given this variability, appropriate guidance on extended glove use for HCWs may require testing to evaluate whether locally available glove options are compatible with locally available disinfectants. This study provides glove testing protocols that could reasonably be done by non-experts in a low-resource setting. Extended use of medical examination gloves may reflect a lack of access to resources, even under a non-pandemic scenario. Disinfection of gloves is not meant to address these systemic health system inadequacies, which demand sustainable supply chains, enhanced equity in health financing, and a view of public health as a global good.

## Data Availability

All data used in the manuscript is publicly available through the listed Google Drive link.

https://docs.google.com/spreadsheets/d/1l2fHxBMUeu3-IV1UKYKopOmCNdChkZMBrCvM0FX-Fbw/edit#gid=1438568418

## Acknowledgements

We thank Andres Ruiz, Kunal Sahasrabuddhe, Salma Elmallah, Adam Gsellman, Matrix Shimizu, Tyler LeBoa, and all others who volunteered their time and energy as glove-testers. We also thank Brian Chang, Gabriel Goldfien, Kimberly Ryan, and Louis Yu for providing safe testing locations despite physical distancing restrictions. Lastly, but not least, we thank Samuel A. Mansfield, Jonathan Patterson, and Juan Cuadros Olave at the University of Houston, Clear Lake, for their intellectual contributions to our work.

## Supplementary Material

### Water Volume Calibration Procedure

The purpose of the calibration test was to determine the baseline volume of water (L) at which 27 punctures resulted in visually detectable leaks. To avoid puncturing through both layers of the glove, we first inserted a dry sponge into the glove and used a permanent marker to clearly mark the location of each hole. Then, using a 30-gauge needle, we poked holes in ten of each glove type and size. We located five holes in each digit and two in the palm of each glove.

Prepared gloves were attached to the PVC water column using a PVC coupling. Beginning with 1L initially, water was slowly added in increments of 0.5L until all leaks were detected in all 27 holes, or until the glove burst. Following the addition of each 0.5 L of water, the tester checked for the presence of leakage at all 27 holes. To spot leaks, a paper towel was used to blot marked holes. We checked for bubbles or streams of water and did not squeeze the glove. Once all 27 holes were leaking or once the glove burst, whichever occurred first, the tester recorded the volume of water that had been added. If the glove burst, we recorded the volume of the prior increment. The final calibration volume was the maximum volume recorded from the ten tests for each glove brand/type.

**Supplementary Table 1:** Decontamination guidelines and application methods.

| Sanitizing agent | Current crisis capacity strategy recommendations (U.S. CDC, updated 23 December 2020) | Selected application method |
| --- | --- | --- |
| Soap and water<br>4,15,18 | Up to ten applications (CDC)<br>No evidence cited | 1. Turn on faucet and wet gloved hands with lukewarm water<br>2. Apply 1 pump of soap to palm of gloved hand<br>3. Rub hands palm-to-palm, spreading the soap and water<br>4. Rub each palm over the back of its opposite hand with interlaced fingers<br>5. Interlock fingers, palm to palm, rubbing them together<br>6. Ball each hand into a fist, twisting its knuckles into the opposite palm<br>7. Wrap and twist each thumb inside the opposite hand's fingers<br>8. Rub the fingertips of each hand on the opposite hand's palm<br>9. Rinse gloved hands with lukewarm water<br>10. Dry hands thoroughly with single-use towel<br>11. Turn off the tap and/or faucet with a towel<br>Duration: 40-60 seconds. |
| ABHR 4,6,15,18 | Up to six applications (CDC) | 1. Apply a palmful of ABHR to one hand |
|  | six applications of ABHR on latex and nitrile gloves resulted in minimal change in glove tensile properties | 2. Rub hands together, palm-to-palm, ensuring even spread of the product on each palm<br>3. Place one palm over the top of the opposite hand, interlocking and moving fingers; repeat by reversing the position of each hand<br>4. Interlock and then fingers, with palms touching one another<br>5. Ball one hand partway into a fist, rubbing knuckles into the opposite hand's palm in a twisting motion; repeat for each hand<br>6. Sanitize the thumbs, wrapping each thumb inside the opposite hand's fingers; rotate fingers around the thumb until completion<br>7. Rub the fingertips — including the thumb — of each cupped hand on the opposite hand's palm in a twisting pattern<br>8. Once hands are dry, the process is complete.<br>Duration: 20-30 seconds. |
| 0.10% sodium hypochlorite solution <sup>4,77</sup> | Up to ten applications (CDC)<br>Manufacturer's testing of Kimberly Clark nitrile gloves found no detectable permeation of 10-13% sodium hypochlorite solution using ASTM standard test methods for permeation after 480 minutes of continuous contact | 1. While gloves are donned, dip hands into a dilute bleach solution for five seconds to ensure complete coverage. Solution should not touch the skin.<br>2. Allow the dilute bleach solution to remain on the donned gloves for one minute (starting after removing gloved hands from the solution) to ensure adequate decontamination. Leave hands in a downward position to reduce the risk of the bleach solution dripping onto arms.<br>3. Rinse dilute bleach solution off gloved hands using water.<br>4. Wipe gloves dry with a clean, absorbent material.<br>5. Check gloves again for signs of damage (e.g., holes, rips, tearing) or degradation (e.g., brittle, stiff, discoloration, tackiness). If damage or degradation is observed, discontinue use and discard the gloves. |

**Supplemental Table 2:** Total recorded leaks by glove location, (n=207)

| Leak location | Recorded leaks n (%)<br>(N=207) |
| --- | --- |
| Interdigital webs | 71 (35%) |
| Palm | 49 (24%) |
| Fingertips | 9 (4.4%) |
| Rest of fingers | 61 (30%) |
| Burst | 9 (4.4%) |
| Undetermined | 7 (3.4%) |

**Supplementary Figure 1:**
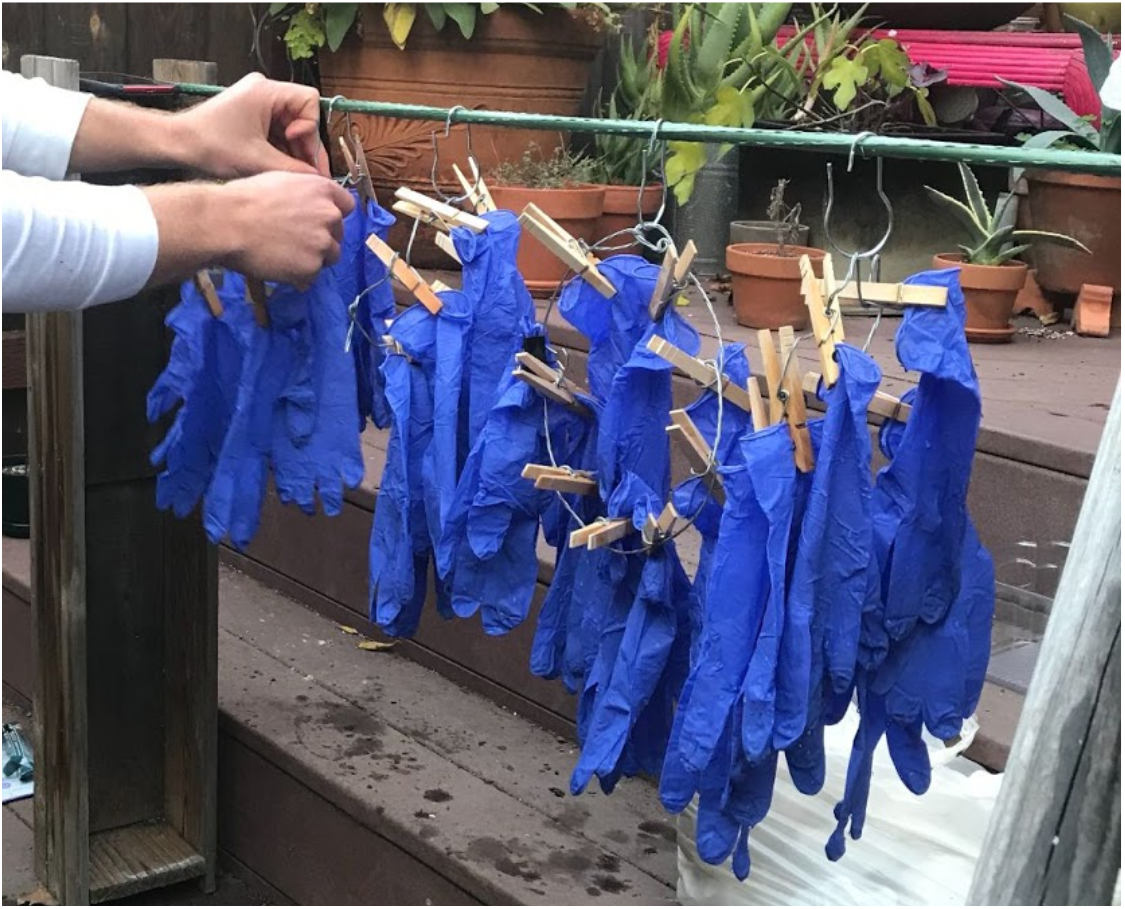
Dilute bleach testing rack. Figure note: Rack used for dipping gloves in bleach solution

**Supplementary Figure 2.**
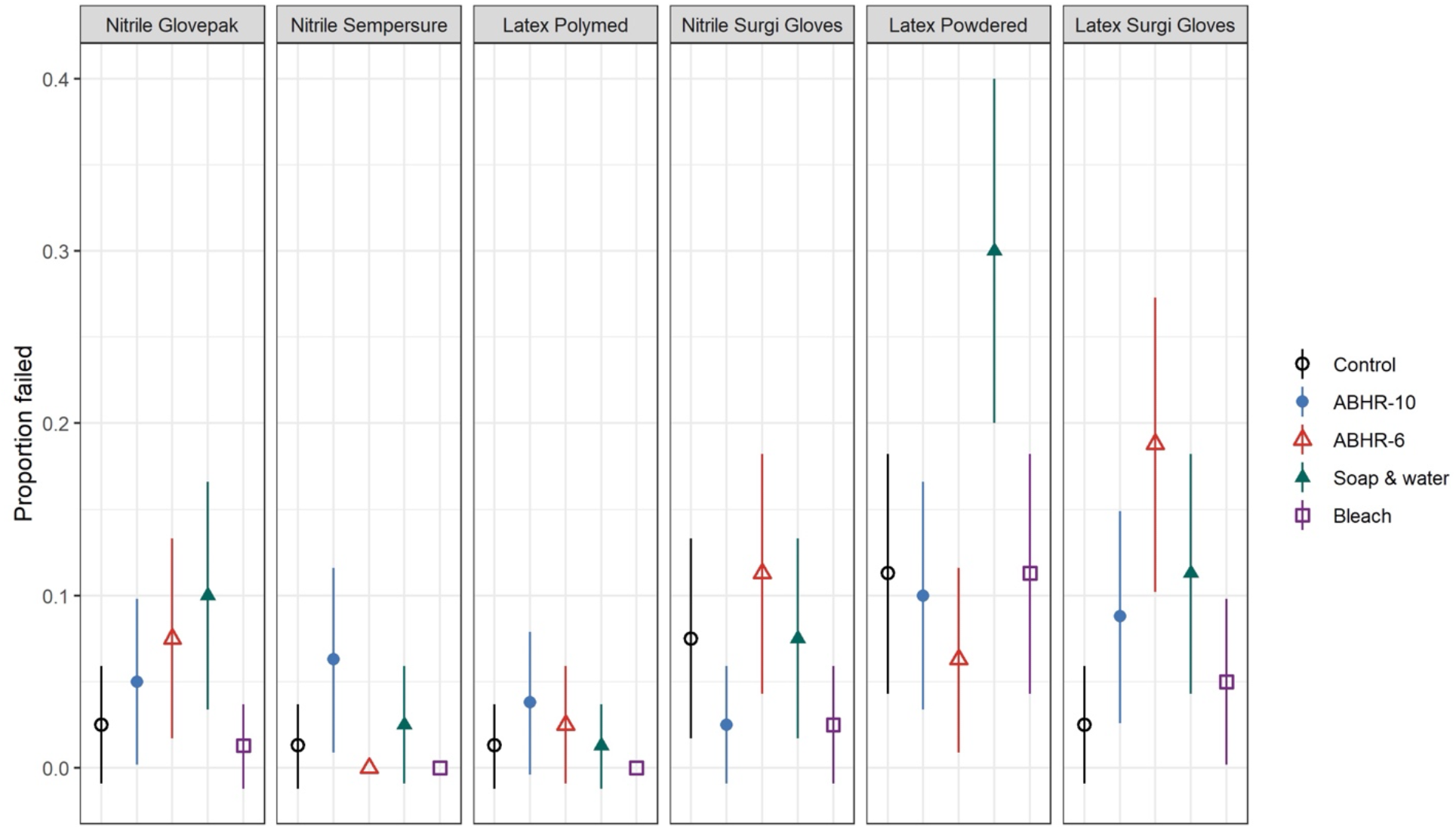
Proportion of gloves failed in each exposure. Figure notes: The left three panels show gloves purchased in the U.S. The right three panels show gloves purchased in India. Each point represents the proportion failed, and the bar represents the 95% confidence interval around the proportion; n=80 for each arm Exposures: ABHR-10 is 10 repetitions of cleansing with alcohol based hand sanitizer; ABHR-6 is 6 repetitions; Soap & Water is 10 repetitions of handwashing with soap and water; Bleach is 10 repetitions of cleansing with dilute bleach solution; Control is gloves out of the box with no exposure

**Supplementary Figure 3.**
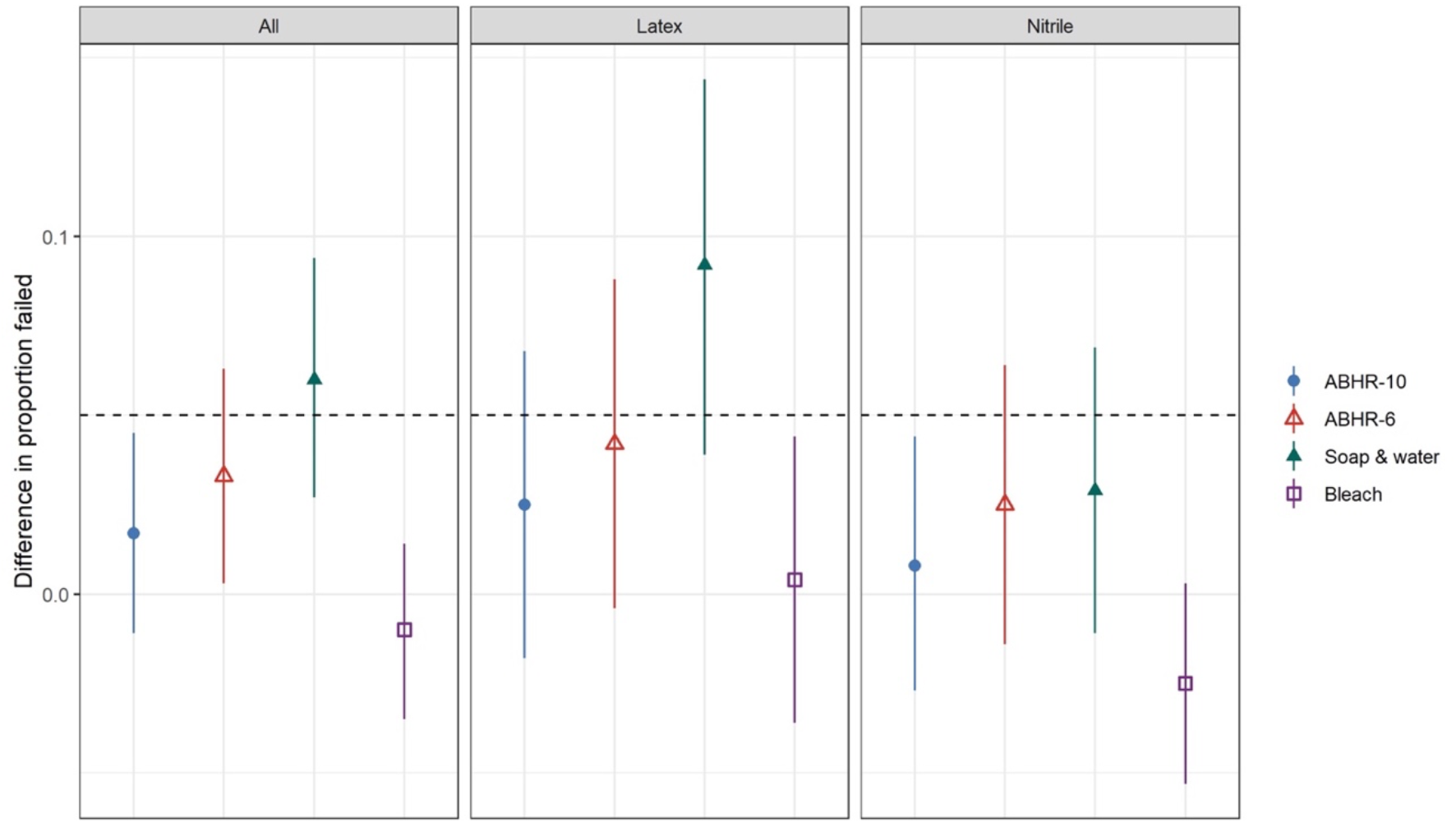
Aggregated differences in proportion failed by exposure and glove material (including powdered gloves) Figure notes: Water-leak test results are shown as differences in the proportion of glove failures between treatment and control (treatment minus control), including the latex powdered gloves. The bar represents the 95% confidence interval for the difference. The dashed line represents the non-inferiority margin (0.05). 6 glove types included; N=480 for all arms. Exposures: ABHR-10 is 10 repetitions of cleansing with alcohol based hand sanitizer; ABHR-6 is 6 repetitions; Soap & Water is 10 repetitions of handwashing with soap and water; Bleach is 10 repetitions of cleansing with dilute bleach solution

